# Patients treated with anti-CD20 therapy can mount robust T cell responses to mRNA-based COVID-19 vaccines

**DOI:** 10.1101/2021.07.21.21260928

**Authors:** Natacha Madelon, Kim Lauper, Gautier Breville, Irène Sabater Royo, Rachel Goldstein, Diego O. Andrey, Alba Grifoni, Alessandro Sette, Claire-Anne Siegrist, Axel Finckh, Patrice H. Lalive, Arnaud M. Didierlaurent, Christiane S. Eberhardt

**Affiliations:** Center for vaccinology, Department of Pathology and Immunology, University Hospital of Geneva & Faculty of Medicine, University of Geneva, Geneva, Switzerland; Department of Medicine, Division of Rheumatology, University Hospital of Geneva & Faculty of Medicine, University of Geneva, Geneva, Switzerland; Department of Neurosciences, Division of Neurology, University Hospital of Geneva & Faculty of Medicine, University of Geneva, Geneva, Switzerland; Department of Diagnostics, Division of Laboratory Medicine; University Hospital of Geneva & Faculty of Medicine, University of Geneva, Geneva, Switzerland; Center for Infectious Disease and Vaccine Research, La Jolla Institute for Immunology, University of California, San Diego, La Jolla, USA; Department of Medicine, Division of Infectious Diseases and Global Public Health, University of California, San Diego, La Jolla, USA; Center for Vaccinology and Department for Paediatrics, Geneva University Hospitals, Emory University School of Medicine, Atlanta, USA; Emory Vaccine Center, Emory University School of Medicine, Atlanta, USA

## Abstract

Patients treated with anti-CD20 therapy are particularly at risk of developing severe COVID-19, however little is known regarding COVID-19 vaccine effectiveness in this population. This study assesses humoral and T-cell responses to mRNA-based COVID-19 vaccines in patients treated with rituximab for rheumatic diseases or ocrelizumab for multiple sclerosis (n=37), compared to immunocompetent individuals (n=22). SARS-CoV-2-specific antibodies were detectable in only 69.4% of patients and at levels that were significantly lower compared to controls who all seroconverted. In contrast to antibodies, Spike (S)-specific CD4+ T cells were equally detected in immunocompetent and anti-CD20 treated patients (85-90%) and mostly of a Th1 phenotype. Response rates of S-specific CD8^+^ T cells were higher in ocrelizumab (96.2%) and rituximab-treated patients (81.8%) as compared to controls (66.7%). Vaccine-specific CD4^+^ and CD8^+^ T cells were polyfunctional but expressed more IL-2 in patients than in controls. In summary, our study suggests that patients on anti-CD20 treatment are able to mount potent T-cell responses to mRNA COVID-19 vaccines, despite impaired humoral responses. This could play an important role in the prevention of severe COVID-19.

## INTRODUCTION

In patients with immune-mediated rheumatic diseases (RD) and multiple sclerosis (MS), immunosuppressive drugs and in particular anti-CD20 therapy are associated with an increased risk of severe COVID-19 [1-3]. Although generally identified as priority groups for vaccination, these patients were not included in pivotal studies evaluating the efficacy of COVID-19 vaccines and their effectiveness in this population is still unknown. Anti-CD20 treatment depletes B cells and impairs antibody responses to classical vaccines [4-6]. Several studies have now confirmed reduced antibody levels and seroconversion rates in anti-CD20 treated patients following SARS-CoV-2 infection [7] and COVID-19 vaccination, irrespective of the underlying disease [8-11]. Although antibodies are likely to play a critical role in preventing infection, recovery from COVID-19 in patients with X-linked agammaglobulinemia suggests that antibodies are not mandatory to overcome disease [12]. T cells may also be involved in protection against COVID-19 [12-14] and memory T cells are readily detectable several months after infection [15]. As B cell could play a role as antigen-presenting cells to naïve T cells, the question remains as whether B-cell depleted patients could still mount a functional T cell response to COVID-19 vaccines, which may provide some level of protection against severe disease.

The aim of our study was thus to characterise and compare T-cell response to mRNA-based COVID-19 vaccines between patients with rheumatic diseases and multiple sclerosis treated with anti-CD20 therapy and immunocompetent controls.

## RESULTS AND DISCUSSION

In order to assess the effect of B cell depletion on vaccine-induced T cell response, we studied a total of 37 patients treated with either ocrelizumab (n=26) for multiple sclerosis (MS) or rituximab (n=11) for rheumatic diseases (RD) compared to 22 age-matched immunocompetent controls (details see Table 1). Most patients with RD were treated for rheumatoid arthritis (n=7) and had more co-morbidities compared to MS patients or controls. Mean age was balanced between groups. Regarding concomitant medication, 5/11 RD patients received another immunosuppressor (mainly methotrexate or corticosteroids), while all MS patients were treated with ocrelizumab only. Hence, this cohort is likely to reflect the effect of short- to long-term treatment with anti-CD20 on vaccine response in the absence of other major confounding factors, in particular age and concomitant immunosuppressive drugs. The interval between last anti-CD20 treatment and 1^st^ vaccine dose was shorter in patients under ocrelizumab (median 24.9 weeks) than in patients treated with rituximab (median 42 weeks), which explained the absence or low (<2 %) percentage of CD19+ B cells at time of vaccination, especially in ocrelizumab-treated patients (Sup Fig 1). Most patients (34/37, 91.9%) and a majority of controls (15/22, 68.2%) had no history of SARS-COV-2 RT-PCR-confirmed infection (review of medical record) prior to vaccination (Table 1) nor a positive anti-nucleoprotein serology (measured 30 days after vaccination, Fig 1A).

**TABLE 1.**

|  | Healthy controls | MS patients | RD patients |
| --- | --- | --- | --- |
| **n** | 22 | 26 | 11 |
| **Female, n(%)** | 15 (68.2) | 14 ( 53.8) | 7 ( 63.6) |
| **Age, median[IQR]** | 54.5 [43.5, 58.8] | 45.6 [39.8, 52.7] | 58.0 [46.4, 64.6] |
| **Comorbidities\*, n(%)** | 3 (13.6) | 6 ( 23.1) | 7 ( 63.6) |
| **History COVID-19 (RT-PCR), n (%)** | 3 (13.6) | 0 | 2 ( 18.2) |
| **Positive anti-N serology, n(%)** | 4 (18.2) | 1 (3.8) | 0 (0) |
| **Vaccine, n(%)** |  |  |  |
| **BNT162b2** | 5 (22.7) | 3 ( 11.5) | 5 ( 45.5) |
| **mRNA-1273** | 17 (77.3) | 23 ( 88.5) | 6 ( 54.5) |
| **Neurologic disease, n(%)** | - |  | - |
| **PPMS** |  | 3 ( 11.5) |  |
| **RRMS** | - | 21 ( 80.8) | - |
| **SPMS** | - | 2 ( 7.7) | - |
| **Rheumatologic disease, n(%)** | - | - |  |
| **Rheumatoid arthritis** | - | - | 7 ( 63.6) |
| **CTD** | - | - | 3 ( 27.3) |
| **Vasculitis** | - | - | 1 ( 9.1) |
| **Anti-CD20 therapy, n(%)<sup>1</sup>** |  |  |  |
| **Ocrelizumab (600 mg)** | - | 26 (100) | - |
| **Rituximab** | - | - | 11 (100) |
| **500 mg** | - | - | 1 ( 9.1) |
| **1000 mg** | - | - | 5 ( 45.5) |
| **1500 mg** | - | - | 1 ( 9.1) |
| **2000 mg** | - | - | 4 ( 36.4) |
| **Time between last treatment and 1<sup>st</sup> vaccine dose, weeks [IQR]** | - | 24.9 [17.3, 26.4] | 42.0 [30.6, 58.7] |
| **Other treatment, n(%)** | - |  |  |
| **Glucocorticoid** |  | 0 | 2 (18.2) |
| **Methotrexate** | - | 0 | 4 (36.4) |
| **Leflunomide** | - | 0 | 1 (9.1) |

PPMS: primary progressive multiple sclerosis, RRMS: relapsing remitting multiple sclerosis, SPMS: secondary progressive multiple sclerosis, CTD: connective tissue disease, IS: immunosuppressive treatment
\*Comorbidities: chronic lung disease, diabetes, hypertension, obesity, depression, cardiovascular disease
<sup>1</sup>The dose mentioned is the total dose that the subject received in around 2 weeks.
<sup>2</sup>Measured between D0 and D60, if not available between the last dose and D0 and if not available, considered as missing

**Figure 1.**
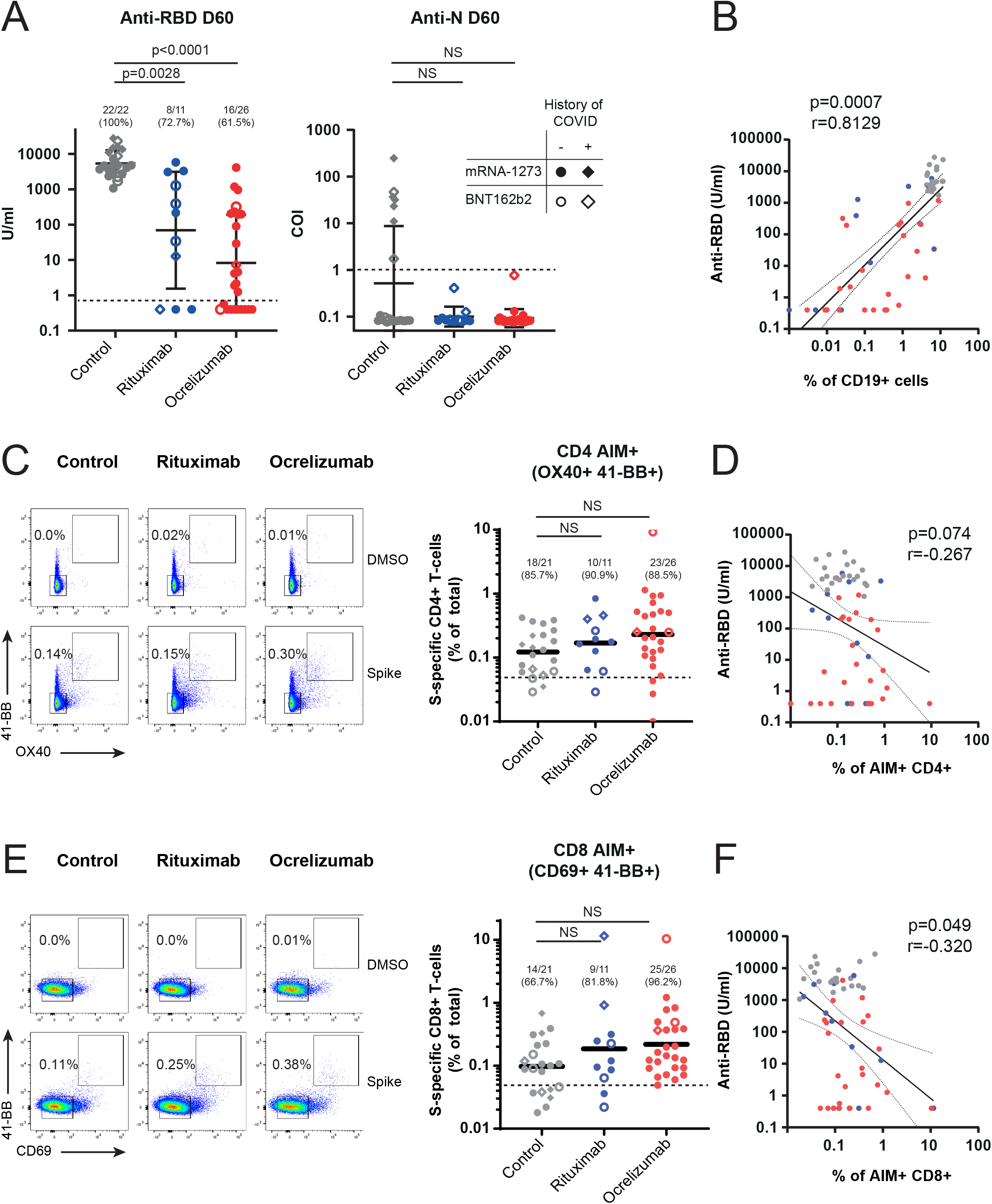
SARS-Cov2 mRNA vaccination induces antigen-specific CD4+ and CD8+ T-cells in Rituximab and Ocrelizumab-treated patients. Levels of anti-SARS-CoV-2 N and RBD total Ig measured in sera of healthy controls (n=22), rituximab (n=11) and ocrelizumab-treated patients (n=26) 30 days after the second dose of BNT162b2 (open symbol) or mRNA-1273 (closed symbol) COVID-19 mRNA vaccines. Dotted line indicates cut-off for seropositivity: anti-RBD; 0.8 U/ml; anti-N: 1 COI. Spearman correlations of anti-RBD antibodies and frequency of CD19+ B-cells in all patients (n=58) (C,E) Representative flow cytometry plots of CD4+ (C) and CD8+ (E) T-cells after PBMC stimulation with DMSO (negative control) and S-peptide pool 30 days after the second vaccination. S-specific AIM+ T-cells are gated as OX40+ 41-BB+ for CD4+ T cells or CD69+ 41-BB+ for CD8+ T-cells (E). Individual data are represented on the right-hand panel with geometric mean. The dotted line represents the limit of detection. Percentages of responders (those with level above limit of detection) are indicated. (D,F) Correlation of anti-RBD total antibodies and AIM+ CD4+ (D) and CD8+ (F) T-cells in all patients (n=58).

Participants were vaccinated either with 2 doses of BNT162b2 (Pfizer/BioNTech; n= 13) or mRNA-1273 (Moderna, n=46) COVID-19 mRNA vaccine at 28 days interval (median 28 days, IQR=0). Immune responses were measured 30 days after the second dose and in a subset of participants (n=20) at time of first vaccination (Sup Fig 2). Both mRNA vaccines are able to elicit antibody and T cell responses in healthy individuals [16, 17]. As recently reported by others [10, 11, 18] and as expected from experience with other vaccines, the level of antibodies specific to the anti-receptor binding domain (RBD) of the SARS-CoV-2 spike (S) protein was significantly lower in both anti-CD20 treated patient populations as compared to controls, irrespective of the vaccine used (geometric mean: 5371 U/ml in controls, 69.3 U/ml in rituximab- and 8.3 U/ml in ocrelizumab-treated patient, Fig 1A). While all controls had seroconverted 30 days after vaccination and regardless of their history of COVID-19, significantly less anti-CD20 treated patients had detectable anti-RBD antibodies (p=001), with a higher seropositivity rate in patients treated with rituximab (8/11; 72.7%) compared to ocrelizumab (16/26; 61.5%). Those RD patients who did not have detectable antibody all received concomitant treatment with methotrexate or corticosteroids. As expected, antibody levels in patients correlated with level of circulating CD19+ B cells (measured at day 30 after vaccination, Fig 1B), which were lower in patients on ocrelizumab due to a more recent treatment. There was no correlation between age and antibody response. The three patients with a known history of COVID-19 or with detectable anti-N antibodies did not have higher antibody responses as compared to those unexposed (Fig 1A, Sup Fig 2A). This suggests that on anti-CD20 treatment, previous exposure to SARS-CoV-2 does not provide an advantage in terms of humoral vaccine response, in contrast to what we (Fig 1A) and others [19] observed in immunocompetent individuals.

T-cell immunity against SARS-CoV-2 is thought to play a role in protection against severe disease[14] and may thus provide for patients under anti-CD20 treatment some level of protection despite their limited antibody response. The number and functionality of T cells is generally maintained after treatment with B-cell targeting drugs, although depletion of some CD20+ T cells, an increase in memory and loss of terminally differentiated CD4+ T cells have been reported [20, 21]. To assess if mRNA vaccines could elicit T cell responses in our patient cohort, we stimulated PBMC collected 30 days after the second vaccine dose with a pool of peptides covering the S-protein [22] and identified S-specific T cells using the activation-induced marker (AIM) assay. S-specific OX40+ 41-BB+ CD4+ T cells were equally induced in immunocompetent and anti-CD20 treated patients (Fig 2C), with a high frequency of responders (85-91%). S-specific CD69+ 41BB+ CD8+ T cells were detectable at similar level in all groups, however there was a statistically significant higher response rate found in ocrelizumab- (96.2%; 25/26) and rituximab-treated patients (81.8%; 9/11) compared to controls (66.7%; 14/21, p=0.02, Fig 2E). Previous history of COVID-19 and the type of mRNA vaccine had no impact on the level of vaccine-specific T-cells. Interestingly, the magnitude of S-specific CD8+ T cells correlated inversely with anti-RBD antibody responses considering all participants (Fig 2F, not significant for CD4+ T cells, Fig 2D). Lastly, the higher frequency of patients with AIM+ CD8+ T cells compared to controls was probably not due to higher levels of pre-existing cross-reactive T cells: in a subset of previously uninfected patients (n=13), S-specific AIM+ CD4+ and CD8+ T cells were undetectable at time of first vaccination (Sup Fig 2B, C).

**Figure 2.**
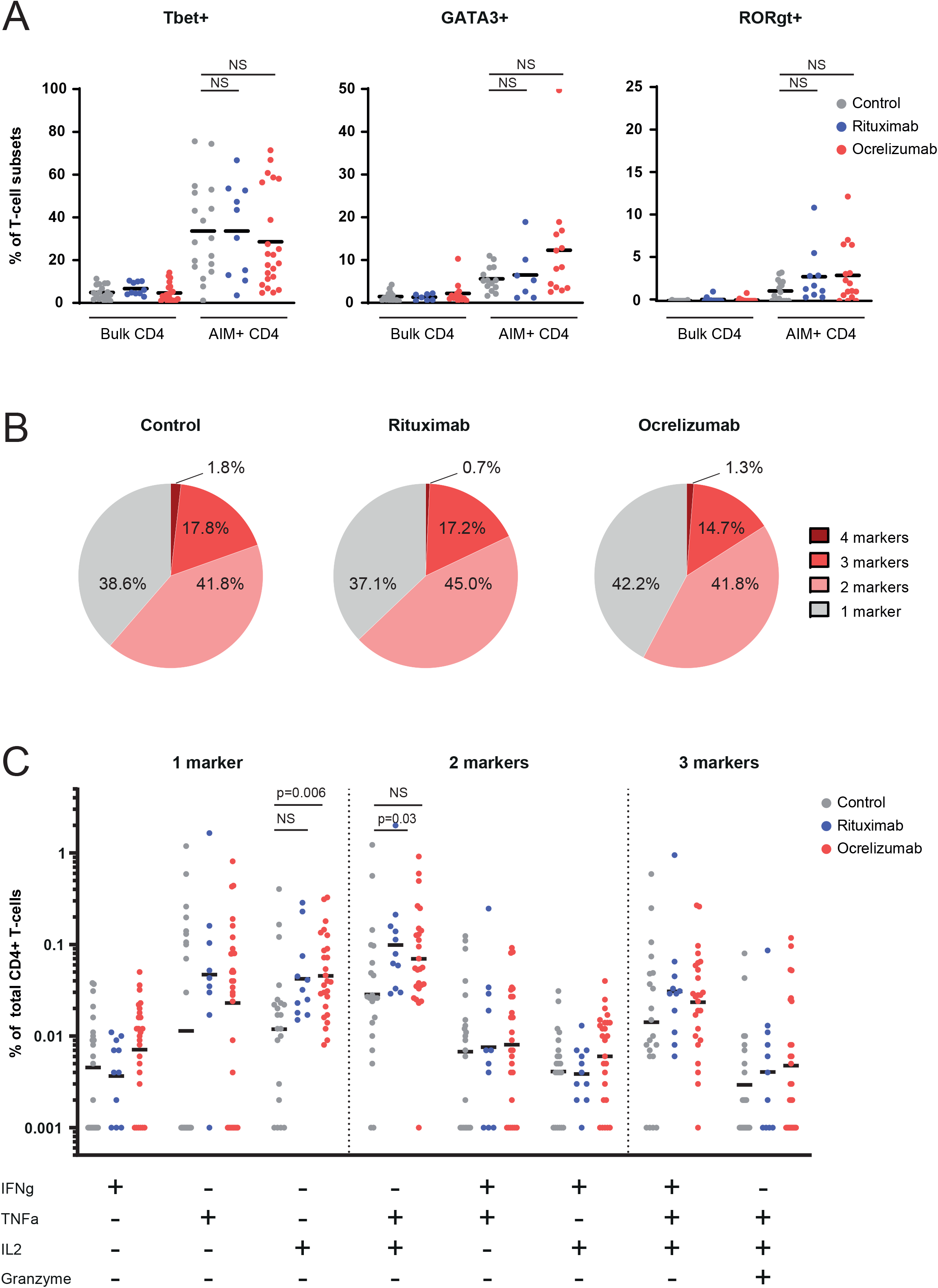
S-specific CD4+ T cell vaccine response is polyfunctional in patients treated with anti-CD20 and produce more IL-2 compared to controls. (A) Expression of Tbet, GATA3, and RORgt in non-specific CD4+ T-cells (“bulk”) and AIM+ S-specific CD4+ T-cells of healthy controls (n=14-18), rituximab-treated patients (n=7-10), and ocrelizumab-treated patients (n=13-22) after stimulation with peptide pool. Analyses were restricted to individuals with detectable AIM+ CD4+ T-cells. (B) Pie chart showing polyfunctionality of S-specific CD4+ T cells of healthy controls (n=21), rituximab-treated patients (n=11) and ocrelizumab-treated patients (n=26). The proportions of CD4+ T-cells expressing 1, 2, 3, or 4 of the activation markers IL2, TNF-a, IFN-gamma or Granzyme B after peptide pool stimulation are shown. (C) Individual data of S-specific CD4+ T-cells expressing different combination of markers (in % of total CD4+ T cells, background subtracted).

This suggests that the benefit of vaccination in terms of T cell responses might be higher in patients with anti-CD20 treatment than in the general population.

We then assessed the functionality of antigen-specific T cells to understand if the quality of T cell responses is altered in the absence of B cells. mRNA COVID-19 vaccines are known to predominantly induced Th1 CD4+ T cells expressing IL-2, IFN-gamma and the transcription factor Tbet rather than Th2 (IL-13+, GATA3+) or Th17 (IL-17+, RORgammaT+) cells [17]. First, we confirmed that a majority of S-specific AIM+ CD4+ T cells expressed Tbet in all patients and controls and found that some ocrelizumab-treated patients had a higher percentage of GATA-3+ T cells, however not reaching statistical significance at the group level (Fig 2A).

Next, we used intracellular cytokine staining to evaluate if vaccine-specific T cells express several cytokines, given that polyfunctional T cells are often associated with improved vaccine-induced protection to viral infection. We found anti-CD20 treated patients had similar level of S-specific CD4+ T cells expressing at least 2 of the markers IL-2, TNF-alpha, IFN-gamma or granzyme B as compared to controls, suggesting a similar polyfunctionality (Fig 2B). The frequency of S-specific CD4+ T cells producing IL-2 and IL-2+TNF-alpha+ in both ocrelizumab and rituximab-treated patients was however higher as compared to immunocompetent controls, while the percentage of CD4+ T cells expressing IFN-gamma alone or in combination with other cytokines was similar (Fig 2C and Sup Fig 3A). There were no detectable IL-13 or IL-17-expressing CD4+ T cells (Sup Fig 3A). Some cytokine-expressing CD4+ T cells co-expressed granzyme B, suggesting that those cells could also have cytotoxic capacity.

In general, S-specific CD8+ T cells expressing either IL-2 or IFN-gamma were detected in more patients treated with anti-CD20 than in controls (Fig 3A). The percentage of IL-2-expressing cells was significantly higher (p=0.013) in Ocrelizumab-treated patient as compared to controls while only a trend was observed for IFN-gamma (p=0.07) and for rituximab-treated patients for all two cytokines. Similar to CD4+ T cells, patients on anti-CD20 had polyfunctional vaccine-specific CD8+ T cells, with a trend for more cells expressing at least 3 markers and significantly more single IL2+ than controls (Fig 3B, C). In general, a higher frequency of S-specific CD8+ T cells co-expressing granzyme and cytokines were found in patients as compared to controls.

**Figure 3.**
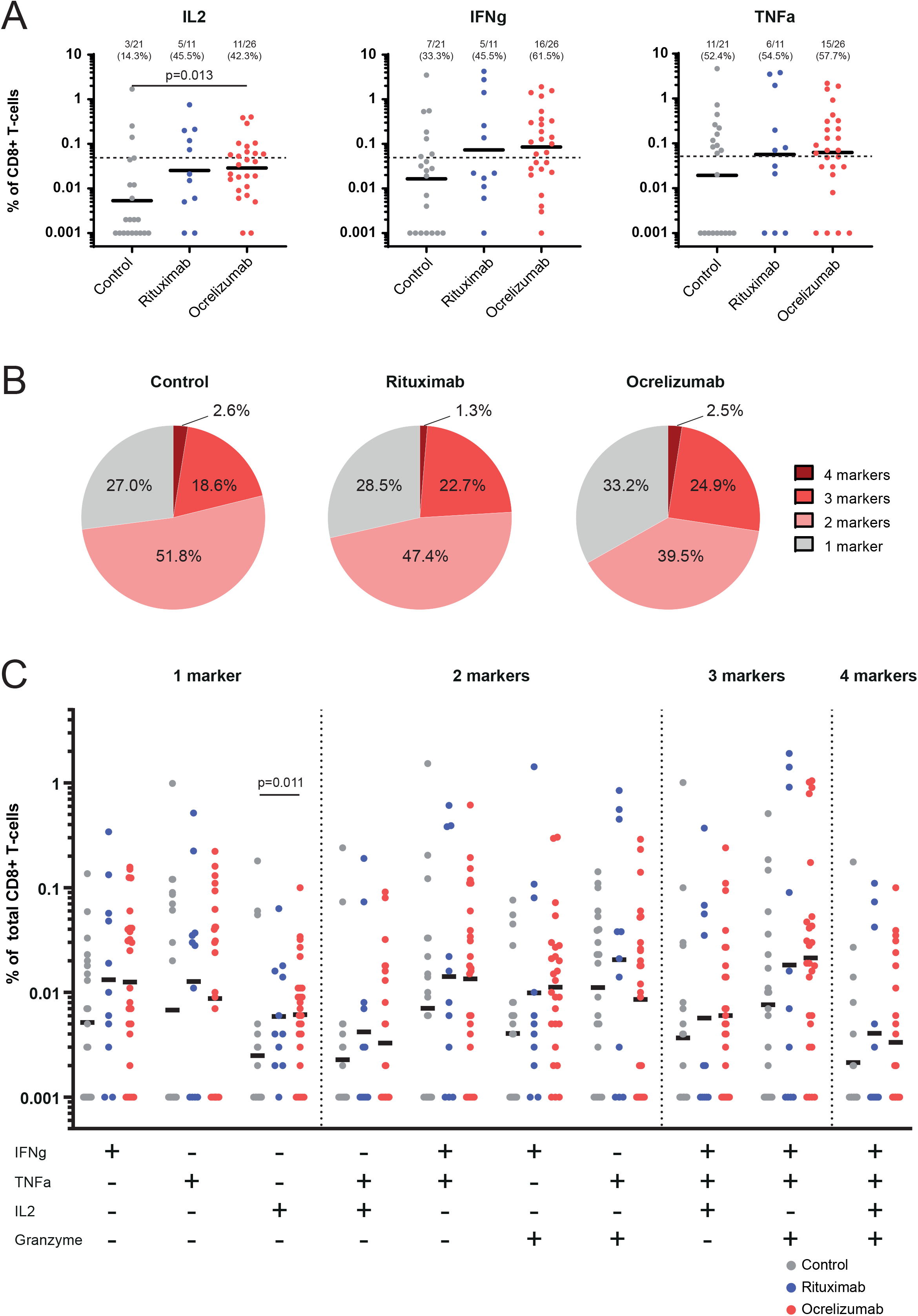
S-specific CD8+ T cell vaccine response are more activated in patients treated with anti-CD20 as compared to controls. (A) Expression of S-specific CD8+ T-cells expressing IFNg, IL-2 or Granzyme B in healthy controls (n=21), rituximab-treated patients (n=11) and ocrelizumab-treated patients (n=26) upon stimulation with peptide pool (background subtracted). (B) Pie chart showing polyfunctionality of S-specific CD8+ T cells shown as proportions of CD8+ T-cells expressing 1, 2, 3, or 4 of the activation markers IL2, TNFa, IFNg or Granzyme B after peptide pool stimulation (C) Individual data of S-specific CD8+ T-cells expressing different combination of markers (in % of total CD8+ T cells, background subtracted).

Altogether, our data suggest that S-specific T cells induced by mRNA vaccines have a similar functional profile but a more activated phenotype in anti-CD20 treated patients as compared to controls. Our finding is consistent with preserved T cell function in these patients and earlier report of cellular responses induced by other vaccines [23]. One hypothesis to explain higher T cell activation in those patients could be the presence of more activated APCs (eg. monocytes) at time of vaccination as a result of B cell depletion [24].

Finally, we assessed the memory phenotype of S-specific AIM+ CD4+ and CD8+ T cells and did not find any difference between groups (Sup Fig 3 and 4). CD8+ T cells had predominantly an effector memory phenotype (CD45RA-CCR7-) while CD4+ T cell phenotypes were equally effector (CD45RA-CCR7-) and central memory (CD45RA-CCR7+). This suggest that in addition to generating polyfunctional T cells, it is likely that mRNA vaccines can generate long-lasting memory T cells in patients treated with anti-CD20. This will be confirmed in follow-up studies looking at the persistence of the cellular response in these patients at 6- and 12-months post vaccination. Interestingly, in hematologic cancer patients treated with anti-CD20, a greater number of CD8 T cells is associated with improved survival to COVID-19, despite impairment in humoral immunity, and 77% of patients had detectable SARS-CoV-2-specific T cell responses [25].

Limitations of our study include the small sample size and the short follow-up after vaccination. We also were not able to correlate T cell findings with clinical protection as the study was not designed to measure efficacy, which is pivotal in the identification of vaccine-responders and the indication for a potential third vaccine dose. Strengths of our study is the prospective design and the ability to evaluate two patient populations under anti-CD20 therapy who have different underlying conditions that do both not greatly affect other axes of immune responses, such as in poly-immunosuppressed patients, or those suffering from lymphoma or leukemia.

In summary, our study suggests that patients with anti-CD20 treatment are able to mount potent T-cell responses to mRNA COVID-19 vaccines similar to immunocompetent controls. Although patients treated with anti-CD20 treatment have decreased humoral responses to mRNA COVID-19 vaccines, elicited T-cell memory response could reduce complications of SARS-CoV-2 infection in this vulnerable population.

## METHODS

### Study participants

We included subjects that were ≥ 18 years of age scheduled to receive the COVID-19 vaccine or having received ≤ 2 COVID-19 vaccine doses in the last 5 weeks. Subjects with SARS-CoV-2 documented infection less than 3 months previous inclusion or ongoing signs of febrile or non-febrile infection were excluded. Further details on the study are in supplementary methods.

### Immunological read-outs

Antibodies were measured using the Elecsys platform (Roche Diagnostics) and seroconversion was defined as > 0.8 IU/ml. For the AIM assay, cells were stimulated overnight with SARS-CoV-2 megapool peptides (by 10 overlapping 15 mers [22]) or in DMSO (negative controls). For intracellular cytokine production, Brefeldin A (Golgiplug, BD) was added to the culture overnight. Cells were stained with specific antibodies and analysed by flow cytometry (see supplementary methods for further details). Percentage of S-specific T cells, AIM+, cytokines+ or granzyme B + cells were calculated by subtracting the value of the corresponding DMSO stimulation control samples.

### Statistics

Clinical data are presented as medians with IQRs for continuous variables and frequency with percentage for categorical variables. For comparison of immune responses between groups, Kruskal-Wallis one-way ANOVA with Dunn’s multiple comparisons was used. Categorical variables were compared using Fisher’s exact test. Correlation analyses were performed using Spearman test. P values from correlations were corrected for multiple comparisons using the False Discovery Rate method.

### Study approval

This prospective observational study was conducted at the Geneva University Hospitals (HUG), Switzerland according to the principles of Good Clinical Practice and was approved by the Geneva Cantonal Ethics Commission (2021-00430). Informed consent was obtained from all participants.

## Data Availability

All data are available in text, figures or as supplementary material. Raw data files (anonymized data)included in this manuscript may be available upon request to the authors.

## AUTHOR CONTRIBUTIONS

CAS, AF, PL, AD and CE conceived the study and wrote the clinical protocol. NM and ISR conducted and analyzed the T cell assays. KL, GB and RG analyzed the clinical data. DA conducted serology testing. RG organized participants recruitment and the collection of clinical specimens. AG and AS provided peptide pools and technical support. NM, AD and CE prepared figures and wrote the manuscript. All authors provided intellectual input and reviewed the manuscript.

## ACKNOWLEDGMENTS

We are grateful to all volunteers for their participation in the study. We thank Mélanie Jaquet, Sandra Fernandes Oliveira, Olivia Studer, Sandrine Bastard and colleagues at the Clinical Research Center, University Hospital and Faculty of Medicine, Geneva for their involvement in patient recruitment and sample collection; Chantal Tougne, Paola Fontannaz, Loïck Staudenmann and Yves Donati for their contributions to processing the clinical samples and support in experimental work. We thank all colleagues that have provided support to participant recruitment, sample collection and experimental work. This work was supported by the HUG private Foundation (CSE, CAS, AD), the NIH NIAID contract no. 75N9301900065 (AS) and by the Giorgi-Cavaglieri foundation (AD).

## CONFLICT OF INTEREST

AS is a consultant for Gritstone, Flow Pharma, CellCarta, Arcturus, Oxfordimmunotech, and Avalia. AD is consultant for Speranza. All of the other authors declare no competing interests. LJI has filed for patent protection for various aspects of vaccine design and identification of specific epitopes.

## Notes

### Competing Interest Statement

A.S. is a consultant for Gritstone, Flow Pharma, CellCarta, Arcturus, Oxfordimmunotech, and Avalia. A.D. is consultant for Speranza. All of the other authors declare no competing interests. LJI has filed for patent protection for various aspects of vaccine design and identification of specific epitopes.

### Funding Statement

This work was supported by the HUG private Foundation, the NIH NIAID contract no. 75N9301900065 and by the Giorgi-Cavaglieri foundation.

### Author Declarations

This prospective observational study was conducted at the Geneva University Hospitals (HUG), Switzerland according to the principles of Good Clinical Practice and was approved by the Geneva Cantonal Ethics Commission (2021-00430).

## REFERENCES

1. Sparks, J.A., et al., Associations of baseline use of biologic or targeted synthetic DMARDs with COVID-19 severity in rheumatoid arthritis: Results from the COVID-19 Global Rheumatology Alliance physician registry. Ann Rheum Dis, 2021.

2. Sormani, M.P., et al., Disease-Modifying Therapies and Coronavirus Disease 2019 Severity in Multiple Sclerosis. Ann Neurol, 2021. 89(4): p. 780–789.

3. Avouac, J., et al., COVID-19 outcomes in patients with inflammatory rheumatic and musculoskeletal diseases treated with rituximab: a cohort study. Lancet Rheumatol, 2021. 3(6): p. e419–e426.

4. Bar-Or, A., et al., Effect of ocrelizumab on vaccine responses in patients with multiple sclerosis: The VELOCE study. Neurology, 2020. 95(14): p. e1999–e2008.

5. Bingham, C.O., 3rd, et al., Immunization responses in rheumatoid arthritis patients treated with rituximab: results from a controlled clinical trial. Arthritis Rheum, 2010. 62(1): p. 64–74.

6. Westra, J., et al., Rituximab impairs immunoglobulin (Ig)M and IgG (subclass) responses after influenza vaccination in rheumatoid arthritis patients. Clin Exp Immunol, 2014. 178(1): p. 40–7.

7. Zabalza, A., et al., COVID-19 in multiple sclerosis patients: susceptibility, severity risk factors and serological response. Eur J Neurol, 2020.

8. Sattler, A., et al., Impaired humoral and cellular immunity after SARS-CoV2 BNT162b2 (Tozinameran) prime-boost vaccination in kidney transplant recipients. J Clin Invest, 2021.

9. Herishanu, Y., et al., Efficacy of the BNT162b2 mRNA COVID-19 vaccine in patients with chronic lymphocytic leukemia. Blood, 2021. 137(23): p. 3165–3173.

10. Furer, V., et al., Immunogenicity and safety of the BNT162b2 mRNA COVID-19 vaccine in adult patients with autoimmune inflammatory rheumatic diseases and in the general population: a multicentre study. Ann Rheum Dis, 2021.

11. Achiron, A., et al., Humoral immune response to COVID-19 mRNA vaccine in patients with multiple sclerosis treated with high-efficacy disease-modifying therapies. Ther Adv Neurol Disord, 2021. 14: p. 17562864211012835.

12. Soresina, A., et al., Two X-linked agammaglobulinemia patients develop pneumonia as COVID-19 manifestation but recover. Pediatr Allergy Immunol, 2020. 31(5): p. 565–569.

13. Gallais, F., et al., Intrafamilial Exposure to SARS-CoV-2 Associated with Cellular Immune Response without Seroconversion, France. Emerg Infect Dis, 2021. 27(1): p. 113–21.

14. McMahan, K., et al., Correlates of protection against SARS-CoV-2 in rhesus macaques. Nature, 2021. 590(7847): p. 630–634.

15. Peng, Y., et al., Broad and strong memory CD4(+) and CD8(+) T cells induced by SARS-CoV-2 in UK convalescent individuals following COVID-19. Nat Immunol, 2020. 21(11): p. 1336–1345.

16. Jackson, L.A., et al., An mRNA Vaccine against SARS-CoV-2 - Preliminary Report. N Engl J Med, 2020. 383(20): p. 1920–1931.

17. Sahin, U., et al., BNT162b2 vaccine induces neutralizing antibodies and poly-specific T cells in humans. Nature, 2021.

18. Spiera, R.S. Jinich, and D. Jannat-Khah, Rituximab, but not other antirheumatic therapies, is associated with impaired serological response to SARS-CoV-2 vaccination in patients with rheumatic diseases. Ann Rheum Dis, 2021.

19. Mazzoni, A., et al., First-dose mRNA vaccination is sufficient to reactivate immunological memory to SARS-CoV-2 in subjects who have recovered from COVID-19. J Clin Invest, 2021. 131(12).

20. Gingele, S., et al., Ocrelizumab Depletes CD20+ T Cells in Multiple Sclerosis Patients. Cells, 2018. 8(1).

21. Nissimov, N., et al., B cells reappear less mature and more activated after their anti-CD20-mediated depletion in multiple sclerosis. Proc Natl Acad Sci U S A, 2020. 117(41): p. 25690–25699.

22. Grifoni, A., et al., Targets of T Cell Responses to SARS-CoV-2 Coronavirus in Humans with COVID-19 Disease and Unexposed Individuals. Cell, 2020. 181(7): p. 1489-1501.e15.

23. Agarwal, A., et al., Rituximab, anti-CD20, induces in vivo cytokine release but does not impair ex vivo T-cell responses. Am J Transplant, 2004. 4(8): p. 1357–60.

24. Lehmann-Horn, K., et al., Anti-CD20 B-cell depletion enhances monocyte reactivity in neuroimmunological disorders. J Neuroinflammation, 2011. 8: p. 146.

25. Bange, E.M., et al., CD8(+) T cells contribute to survival in patients with COVID-19 and hematologic cancer. Nat Med, 2021.

