## supplementary material for "Patients treated with anti-CD20 therapy can mount robust T cell responses to mRNA-based COVID-19 vaccines"

### SUPPLEMENTARY FIGURES

#### Supplementary Figure 1.

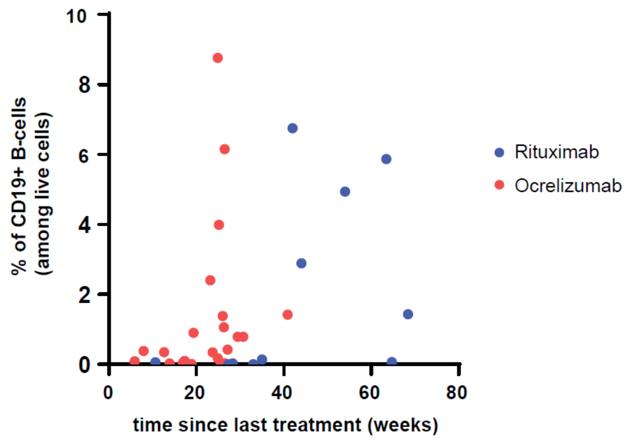


**Supplementary Figure 1. Percentage of CD19+ B cells and time between vaccination and last anti-CD20 treatment**

Y axis: percentage of CD19+ B cells (in total cells) measured by flow cytometry in PBMC collected at day 30 after the second dose in rituximab or ocrelizumab-treated patients.

X axis: corresponding time between the first vaccination (day 0) and the last treatment with anti-CD20 antibody, for each patient, as reported in table 1.

### Supplementary Figure 2

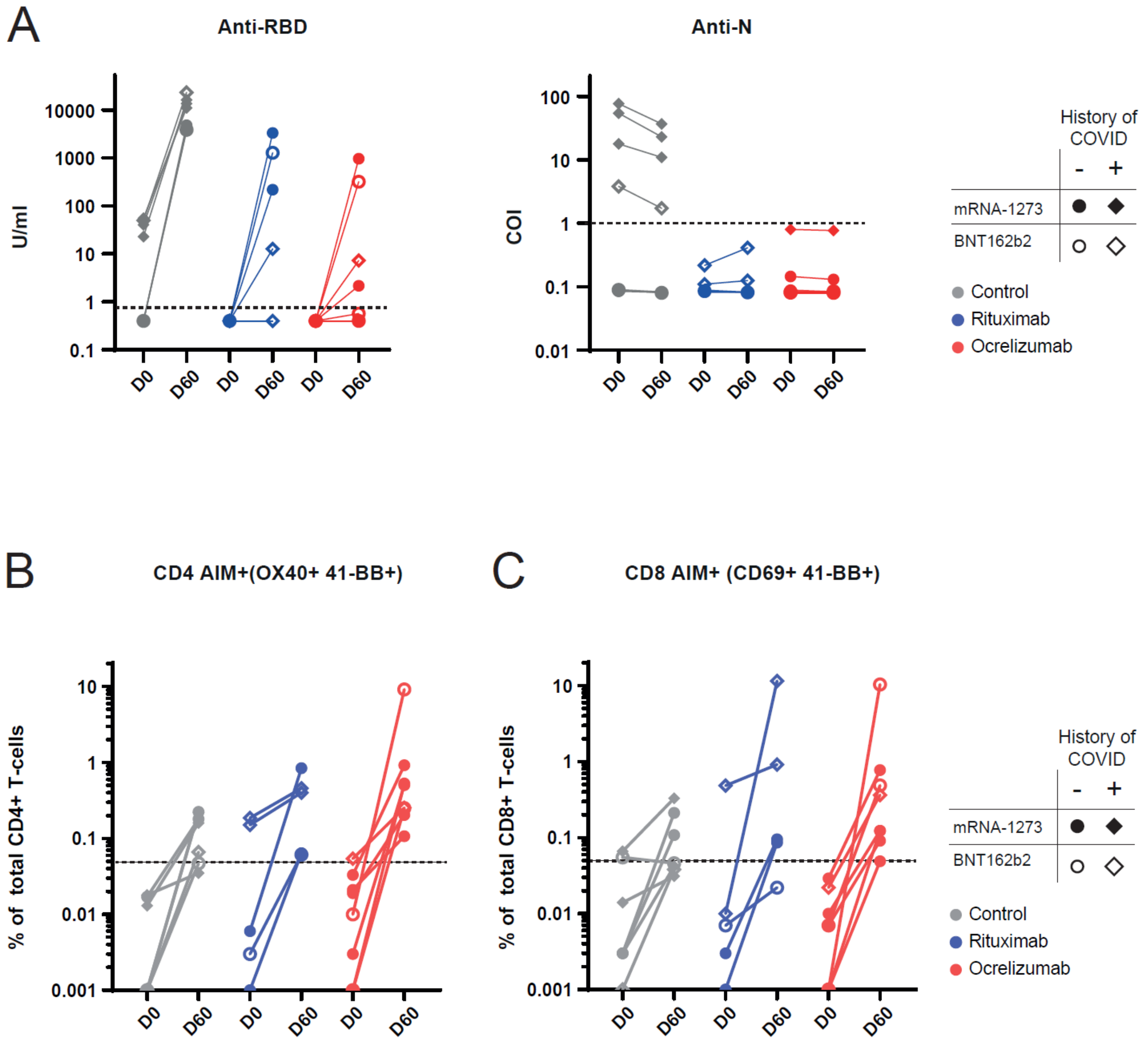


**Supplementary Figure 2. Kinetics of anti-SARS-CoV-2 antibodies and Spike-specific T-cells in healthy controls and anti-CD20-treated patients**

**(A)** Levels of anti-SARS-CoV-2 N and RBD total Ig measured in sera of healthy controls (n=7), rituximab (n=5) and ocrelizumab-treated patients (n=8) before vaccination and 30 days after the second dose of BNT162b2 (open symbol) or mRNA-1273 (closed symbol) COVID-19 mRNA vaccines. Dotted line indicates cut-off for seropositivity: anti-RBD; 0.8 U/ml; anti-N: 1 COI. **(B, C)** Levels of Spike-specific CD4+ T-cells (OX40+ 41-BB+) (**B**) and Spike-specific CD8+ T-cells (CD69+ 41-BB+) (**C**) before vaccination and 30 days after the second dose of BNT162b2 or mRNA-1273 COVID-19 mRNA vaccines in healthy controls (n=7), rituximab (n=5) and ocrelizumab-treated patients (n=8). The dotted line represents the limit of detection.

**Supplementary Figure 3**

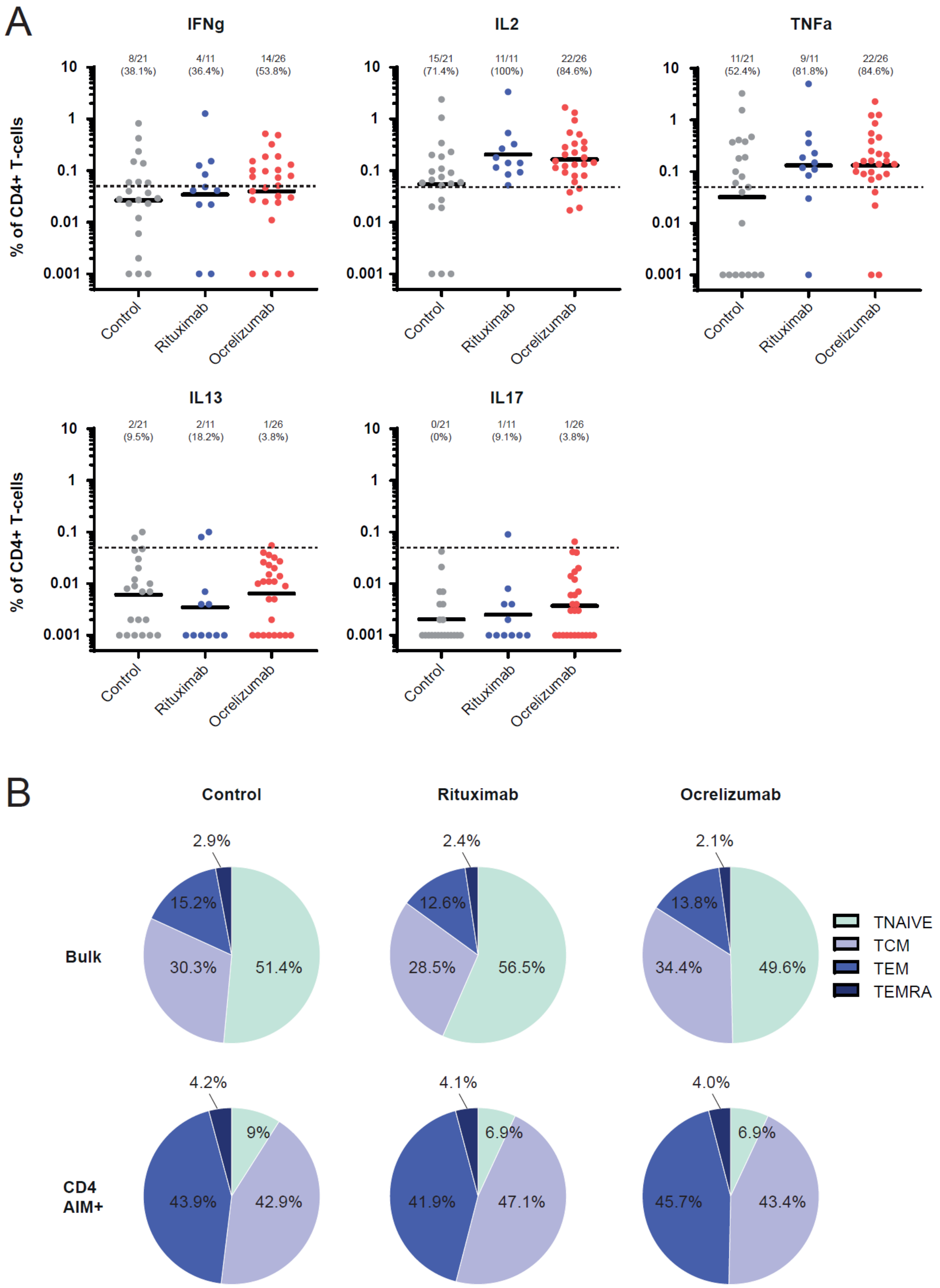


**Supplementary Figure 3. Effector memory phenotype of spike-specific CD4+ T-cells in anti-CD20 treated patients**

**(A)** Expression of S-specific CD4+ T-cells expressing IFNg, IL-2, TNFa, IL13, IL17, or granzyme B in healthy controls (n=21), rituximab-treated patients (n=11) and ocrelizumab-treated patients (n=26) upon stimulation with peptide pool (background subtracted). The dotted line represents the limit of detection. **(B)** Pie charts showing the proportions of memory phenotype of bulk non-specific CD4+ T-cells (AIM- CD4+ T-cells) and Spike-specific CD4+ T-cells (AIM+ CD4+ T-cells) of healthy controls (n=18), rituximab-treated patients (n=10) and ocrelizumab-treated patients (n=23) are shown. The proportions of naïve T-cells ($T_{NAIVE}$, CD45RA+ CCR7+), central memory ($T_{CM}$, CD45RA- CCR7+), effector memory ($T_{EM}$, CD45RA- CCR7-) and $T_{EMRA}$ (CD45RA+ CCR7-) are shown. Analyses were restricted to individuals with detectable AIM+ CD4+ T-cells.

**Supplementary Figure 4**

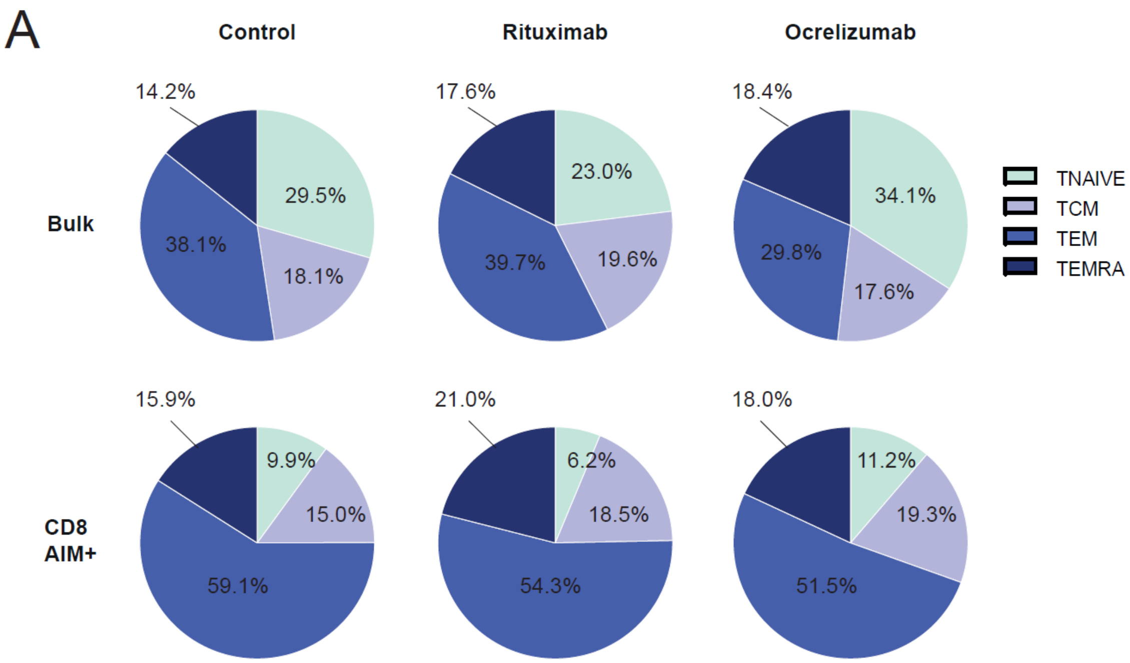


**Supplementary Figure 4. Effector memory phenotype of spike-specific CD8+ T-cells in anti-CD20 treated patients**

**(A)** Pie charts showing the proportions of memory phenotype of bulk non-specific CD8+ T-cells (AIM- CD8+ T-cells) and Spike-specific CD8+ T-cells (AIM+ CD8+ T-cells) of healthy controls (n=14), rituximab-treated patients (n=9) and ocrelizumab-treated patients (n=25) are shown. The proportions of naïve T-cells ($T_{NAIVE}$, CD45RA+ CCR7+), central memory ($T_{CM}$, CD45RA- CCR7+), effector memory ($T_{EM}$, CD45RA- CCR7-) and $T_{EMRA}$ (CD45RA+ CCR7-) are shown. Analyses were restricted to individuals with detectable AIM+ CD8+ T-cells.

### SUPPLEMENTARY METHODS

#### Serum isolation

Whole blood was collected in a 3ml tube (BD, 367957) and centrifuged for 10 minutes at 1000g at 4°C. Serum was collected, aliquoted and stored at -80°C until use.

#### PBMC isolation

Whole blood was collected in CPT tubes (BD, 232782) and human peripheral blood mononuclear cells (PBMC) were isolated by density gradient centrifugation. Whole blood was centrifuged at 1600g for 24 minutes at room temperature. Red blood cells were lysed using ACK lysis buffer (Lonza). Cells were washed, counted, and cryopreserved in FCS containing 10% DMSO in liquid nitrogen until further use.

#### Activation induced marker assay

Cryopreserved PBMC were thawed in complete RPMI supplemented with 10%FCS, essential amino acids ad penicillin/streptomycin in the presence of benzonase (50U/ml). Cells were washed, counted, and resuspended at $20.10^{6}$ cells /ml in complete RPMI. Cells were further rested for 6h and stimulated overnight with 1ug/ml of SARS-Cov2 megapool peptides (CD4-S) in U-bottom 96 well plates at $1\text{-}2.10^{6}$ cells per well at 37°C. Cells were stimulated with SEB at 1ug/ml as positive controls and unstimulated samples with an equimolar volume of DMSO were used as negative controls. SARS-CoV2 megapool peptides were prepared as described previously (10.1016/j.cell.2020.05.015). Cells were then surface stained for 30 minutes at 4°C, fixed/permeabilized using the eBioscience FoxP3 transcription factor buffer kit (Thermofischer scientific), and stained with the Tbet, RORgt, GATA3 transcription factors for 30 minutes at 4°C.

#### Intracellular cytokine staining assay

For the intracellular cytokine staining, PBMC were cultured in the presence of SARS-CoV2 megapool peptides in the presence of Brefeldin A (Golgiplug, BD, 1/1000) overnight at 37°C. Cells were surface stained for 30 minutes at 4C, fixed/permeabilized using cytofix/cytoperm fixation/permeabilization kit (BD, reference 554714) and stained for the intracellular cytokines IFNg, IL2, TNFa, IL17, IL13, granzyme for 30 minutes at 4°C.

Data analysis were performed using FlowJo software (version 10, Treestar). For dead cell exclusion, LIVE/DEAD fixable Aqua kit was used (ThermoFischer). All antibodies were purchased from Biolegend or BD Bioscience and listed in the extended data table 1 below.

**Statistics**

Statistical analysis was performed in GraphPad Prism software (version 8.0.2). One-way ANOVA with Dunn's multiple comparisons was used (unless otherwise stated in the figure legends). Correlation analyses were performed using Spearman test. P values from correlations were corrected for multiple comparisons using the False Discovery Rate method. Categorical variables were compared using Fisher's exact test.

Data presented in linear scale were expressed as mean + Standard Deviation (SD). Data presented in logarithmic scales were expressed as Geometric Mean + Geometric Standard Deviation (SD). The percentages of $AIM^+$ cells or $cytokines^+$ cells obtained after SARS-CoV-2 peptide stimulation have been subtracted with the percentages of positive cells obtained after DMSO stimulation. The limit of detection for activation induced (AIM) and intracellular cytokine staining (ICS) assay was 0.05%.

| **Anticorps** | **Fluorochrome** | **Clone** | **Brand** | **Reference** |
|---|---|---|---|---|
| CD3 | A700 | SK7 | Biolegend | 344822 |
| CD4 | APC-H7 | RPA-T4 | BD | 560158 |
| CD8 | BUV737 | SK1 | BD | 612754 |
| CD45RA | BV785 | HI100 | Biolegend | 304140 |
| CCR7 | PE-Dazzle 594 | G043H7 | Biolegend | 353236 |
| CD69 | PerCP-Cy5.5 | FN50 | BD | 560738 |
| OX40 | BUV395 | ACT35 | BD | 743286 |
| 4-1BB | APC | 4B4-1 | Biolegend | 309810 |
| IFNg | BV421 | 4S.B3 | Biolegend | 502532 |
| TNFa | BV650 | MAb11 | BD | 563418 |
| IL17a | FITC | BL168 | Biolegend | 512304 |
| IL2 | PE | MQ1-17H12 | Biolegend | 500307 |
| Granzyme | PE-Dazzle594 | QA16A02 | Biolegend | 372216 |
| IL13 | PE-Cy7 | JES10-5A2 | Biolegend | 501914 |
| Tbet | BV421 | 4B10 | Biolegend | 644816 |
| RORgt | FITC | Q21-559 | BD | 563621 |
| GATA3 | PE-Cy7 | L50-823 | BD | 560405 |

**Extended Table 1. Antibodies list**
